# Efficacy of Sofosbuvir plus Ledipasvir in Egyptian patients with COVID-19 compared to standard treatment: Randomized controlled trial

**DOI:** 10.1101/2021.05.19.21257429

**Authors:** Mohamed Abdel-Salam Elgohary, Eman Medhat Hasan, Amany Ahmad Ibrahim, Mohamed Farouk Ahmed Abdelsalam, Raafat Zaher Abdel-Rahman, Ashraf Ibrahim Zaki, Mohamed Bakr Elaatar, Mohamed Thabet Elnagar, Mohamed Emam Emam, Mahmoud Moustafa Hamada, Taimour Mohamed Abdel-Hamid, Ahmad Samir Abdel-Hafez, Mohamed G. Seadawy, Ahmad Rashad Fatoh, Mohamed Ali Elsaied, Marwa Abdel-Rahman Sakr, Ahmed Omar Elkady, Mohamed Muawad Shehata, Osama Mohamed Nawar, Mohamed Abu-elnaga Selem, Mohamed Saeed Abd-aal, Hany Hafez Lotfy, Tarek Refaat Elnagdy, Sherine Helmy, Magdy Amin Mubark

## Abstract

**Background:** COVID-19 is a pandemic disease caused by SARS-CoV-2, which is an RNA virus similar to HCV in the replication process. Sofosbuvir/ledipasvir is an approved drug by the FDA to treat HCV infection. This study investigates the efficacy of Sofosbuvir/ledipasvir as a treatment for patients with moderate COVID-19 infection..

**Methods:** This is a single-blinded parallel-randomized controlled trial. The participants were randomized equally into the intervention group received Sofosbuvir/ledipasvir (S.L. group), and the control group received Oseltamivir, Hydroxychloroquine, and Azithromycin (OCH group). The primary outcomes were the cure rate over time and the incidence of serious adverse events. The secondary outcomes included the laboratory findings.

**Results:** Two hundred and fifty patients were divided equally into each group. Both groups were similar regarding gender, but age was higher in the S.L. group (p=0.001). In the S.L. group, 89 (71.2%) patients were cured, while only 51 (40.8%) patients were cured in the OCH group. The cure rate was significantly higher in the S.L. group (RR=1.75, p<0.001). Kaplan-Meir plot showed a considerably higher cure over time in the S.L. group (Log-rank test, p=0.032). There were no deaths in the S.L. group, but there were six deaths (4.8%) in the OCH group (RR=0.08, p=0.013). Seven patients (5.6%) in the S.L. group and six patients (4.8%) in the OCH group were admitted to ICU (RR=1.17, P=0.776). There was no significant difference between treatment groups regarding Total Leukocyte Count, Neutrophils count, Lymph and Urea.

**Conclusion:** Sofosbuvir/ledipasvir is suggestive of being effective in treating patients with moderate COVID-19 infection. Further studies are needed to compare Sofosbuvir/ledipasvir with the new treatment protocols.

## Introduction

COVID-19 (also called coronavirus disease) is a pandemic disease that first appeared in China then spread worldwide and caused 2,836,363 deaths among 130,108,655 infected cases around the world until now (1). The most common symptoms of COVID-19 include fever, cough, shortness of breath, fatigue, muscle or body aches, headache, loss of taste or smell, sore throat, congestion or runny nose, and diarrhea (2). The most common complications include acute respiratory failure, pneumonia, acute respiratory distress syndrome, acute liver, kidney, and cardiac injury (3). COVID-19 is caused by SARS-CoV-2, a beta genus member of the coronavirus (3). Globally, scientists are competing to find drugs to treat COVID-19. Some drugs have been tested in clinical trials quickly and have shown primary efficacy against SARS-CoV-2. Others have been incorporated into a number of guidelines. (4).

SARS-CoV-2 is similar to hepatitis C virus (HCV) in the replication process, as both depend on NS5B RNA-dependent RNA polymerase (NS5B-RdRp) and NS5A, which are essential for the replication process (5, 6). SARS-CoV-2 is also similar to influenza virus in some structural proteins like S protein and Nucleoprotein and non-structural proteins like RNA-directed RNA polymerase (Pol/RdRp), papain-like protease (PLpro), and 3C-like protease (3CLpro) (7).

Sofosbuvir/ledipasvir is an approved drug by the FDA to treat HCV infection. Sofosbuvir causes inhibition of NS5B-RdRp, which is an essential enzyme in the replication process of the HCV virus. On the other hand, ledipasvir inhibits NS5A, necessary for RdRp function (5).

Sofosbuvir/ledipasvir may be beneficial against COVID-19 because SARS-CoV-2 and HCV have similar proteins and enzymes required for the replication process. An experimental study found that Sofosbuvir/ledipasvir is effective against SARS-CoV-2 (6).

Oseltamivir is a neuraminidase inhibitor approved by FDA for influenza (8). Neuraminidase protein is not encoded by SARS-CoV-2 (9). However, Oseltamivir can bind effectively to the active site of key proteins in SARS-CoV-2, which makes it beneficial against COVID-19 (7).

Hydroxychloroquine (HCQ) was proved to have anti-SARS-CoV-2 in vitro (10). A combination of Oseltamivir, Hydroxychloroquine, and Azithromycin was the standard of care in Egypt during the data collection time; however, its use in COVID-19 patients was not beneficial and stopped in the Solidarity Trial (11).

This randomized controlled trial investigates the efficacy of Sofosbuvir/ledipasvir in the treatment of COVID-19 compared to the standard of care.

## Methods

### Study design

This randomized controlled clinical trial is a prospective, comparative, single-blinded (from the patient side), randomized study conducted on 250 patients, divided into two equal groups. The intervention group (S.L. group) received Sofosbuvir/Ledipasvir. In contrast, the control group (OCH group) received the standard of care, Oseltamivir, HCQ, and Azithromycin (the local medical committee of Almaza Fever Hospital guided standard treatment protocol for COVID-19). The Institutional Review Board of Armed Forces College of Medicine approved the study (Date: 12-04-2020, S.N.: 14). The trial is registered at clinicaltrial.gov registry with registration number NCT04530422. The study was conducted by following all the CONSORT checklist 2010 steps (12).

### Participants

The inclusion criteria were pneumonic patients with SARS-COV-2 infection confirmed to be positive by RT-PCR. They demonstrated moderate cases criteria: fever (measured temperature of at least 38 °C), respiratory symptoms (cough, shortness of breath), and imaging-confirmed pneumonia. Inclusion criteria also included age more than 18 and less than 75 years old. Female patients enrolled in the study should have no planned pregnancy for six months, with the administration of proper contraceptive measures within 30 days from the first therapeutic dose of the investigational drugs. Patients agreed to sign an informed consent to participate in the current study and that they would not participate in other clinical trials within 30 days from the last administration of the study drugs.

Exclusion criteria were Mild COVID-19 patients who had mild symptoms without evidence of viral pneumonia or hypoxia and severe COVID-19 patients who met one of the following conditions: (1) Respiratory rate (R.R.) ≥ 30 times/min; (2) SaO2/SpO2 ≤ 93% in resting-state; (3) arterial partial pressure of oxygen (PaO2)/concentration of oxygen (FiO2) ≤ 300 mmHg. Critical COVID-19 patients with one of the following conditions: (1) respiratory failure and need mechanical ventilation; (2) shock; (3) other organ failure combined with ICU treatment; (4) severe liver disease (such as child Pugh score ≥ C, AST > five times upper limit) were excluded. Patients with contraindications specified for any drugs used in the study or who received antiviral eradication therapy for hepatitis C or B viruses within the previous six months were excluded.

Pneumonia was assessed on admission using C.T. Severity Scoring System (CT-SSS) and CO-RADS (Percentage per lobe with a max of 5 points for each lobe and 25 points for both lungs) (13).

COVID-19 RT-PCR test was done by extracting the DNA of the virus by using either device (QIA cube or QIA symphony). Then the process was applied to prepare and implement the RT-PCR step by using chemicals to detect the COVID-19.

### Interventions

Patients assigned to the S.L. group (125 patients) received Sofosbuvir plus Ledipasvir once daily for 15 days, then followed up to day 21..

Patients in the OCH group (125 patients) received Oseltamivir 150 mg q 12 hours for 10 days, HCQ 400 q 12 hours for one day followed by 200mg q 12 hours for nine days, and Azithromycin 500mg one time, followed by 250mg once daily for 6 days.

Additional medications were given, including third-generation cephalosporin Ceftriaxone 2 gm /24 hours for seven days, methylprednisolone 1 mg/kg/day for seven days, and prophylactic low molecular weight heparin (enoxaparin) 40 mg/24 hours was given throughout the hospitalization period.

Patients were evaluated as scheduled on days 0, 5, 10, and 15 days, then up to 21 days for follow up. The evaluation consisted of clinical and laboratory investigations, including C.T. chest.

Serum ferritin and Interleukin 6 levels (IL 6) were asked for patients with suspected cytokine storm (worsened clinical condition, especially fever & dyspnea ± C.T. progression). Selective cytokine blockade (tocilizumab, 400 mg by I.V infusion) was given with evident high IL6 (14).

Medication was stopped at any time if there was any clinical, radiological, or laboratory deterioration.

Any patient demonstrates worsening of symptoms; radiological progression with virological persistence within at least five days of the therapeutic evaluation period of the study after exclusion of cytokine storm was considered as a clinical failure and was shifted to the other management protocol. Treatment was terminated at any time by a multidisciplinary team if a severe side effect occurred, which was attributed to the medications used,e.g., cardiac arrhythmia, deteriorated liver or kidney function, or, unfortunately, the patient died.

### Outcomes

The primary outcomes were the cure rate over time, length of hospital stay and the incidence of serious adverse events that lead to ICU admission or death. The secondary outcomes were the time to virilogical cure as detected by PCR and Chest CT findings.

The outcomes were measured at 0, 5, 10, and 15 days from the first therapeutic dose.

Discharge criteria were resolution of symptoms (Normal body temperature for at least three days and significantly improved respiratory symptoms), Radiological improvement of pneumonic pattern in C.T. chest, and documented virological clearance in two samples least 24 hours apart. Discharge criteria also included no co-morbidities or complications, which require hospitalization and SpO2 >93% without assisted oxygen inhalation.

### Sample size

The null hypothesis is that number of events (cure rate based on clinical status) during phase [up to 15 day] and follow up phase [up to 21 days] in COVID-19 patients, is equal while treated with the combined therapy SOF+Ledi compared to the current MOH regimens. A minimum of 95 subject in each arm was required to fulfill power of 80%. The calculations based on equivalent design as hazard ratio for cure rate as defined by clinical status uptp day 15 on treatment and day 21 follow up in COVID-19 patients, is equal while treated with the combined therapy SOF+Ledi and the current MOH regimen (OCH)

### Randomization

Patients were randomly allocated to one of the two groups. Randomization is applied through computer-generated numbers and concealed using a sequentially numbered sealed opaque envelope.

### Statistical methods

R version 3.5.1 (2018-07-02) -- “Feather Spray” software for windows was used for the statistical analysis. The result is considered significant if it has a p-value lower than 0.05 as an alpha point. Continuous data were described as mean ± standard deviation, while categorical data were expressed as frequency and percentage. An independent t-test was used to compare continuous data, a nd a chi-square test was used to compare categorical data.

Kaplan-Meir plot and log-rank test were used to calculate the cure rate over time. Cox regression was used to adapt for the significant age difference between both groups.

Two way repeated measures ANOVA test was to calculate the change in the laboratory findings over time in each group and the entire sample.

## Results

Two hundred and fifty patients were randomly allocated to two equal groups S.L. group and the OCH group. Each group is formed of 125 COVID-19 positive patients. Patients were recruited from April 15 until the end of June 2020. Six patients in the S.L. group withdrew from the study, while there were no dropouts in the OCH group (Fig 1).

**Figure 1:** Study flowchart

The two groups were similar regarding gender (p=0.113), but the S.L. group was significantly higher than the OCH group regarding age (p=0.001). Fever was the most presenting symptom, and it was none significantly higher in the OCH group (p=1). On the other hand, sore throat was the least presenting symptom, and it was none significantly higher in the OCH group (P=1).

Pneumonia prevalence was higher in the S.L. group (p=0.003). Other clinical and laboratory findings were measured and illustrated in table1.

### Clinical outcomes

In the S.L. group, there are 90 (72%) patients were cured within 14 ± 2 days with median length of stay 16 ± 4 days, while in the OCH group, 50 (40%) patients were cured within 24 ± 14 days with median length of stay 25 ± 8 days (Fig 3). S.L. group was significantly higher than the OCH group (RR= 2.07 with CI: 1.456 - 2.955, p<0.0001).

Kaplan-Meier plot showed that the S.L. group was significantly 2 folds superior regarding the cure rate over time (log-rank=16.98) (Fig 2). Cox regression between the treatment groups was performed considering the significant difference between the two groups regarding other significant covariates at baseline (Fig 4).

**Figure 2:** Kaplan-Meier plot for treatment groups time to clinical cure

**Figure 3:** Kaplan-Meier plot for treatment groups considering the overall patient length of stay.

**Figure 4:** Cox regression between the treatments groups regarding other suspected covariates at baseline.

The forest plot showed that after adjusting for the effect of the covariates that were significant at baseline between the two treatment groups, the adjusted p value was < 0.001 (RR=2.31 with CI: 1.54 – 3.5). Proving that SL group was 2.3 folds higher in the curing rate than OCH group even by taking in consideration the impact of baseline significant covariates.

There were no deaths in the S.L. group, but there were six deaths (4.8%) in the OCH group (RR=0.08, p=0.013). Seven patients (5.6%) in the S.L. group and six patients (4.8%) in the OCH group were admitted to ICU (RR=1.17, P=0.776).

### Laboratory outcomes

Repeated measures ANOVA was used to analyze laboratory findings over the study period within each group and in total.

Total Leukocyte Count tests showed no difference over time; Wilk’s Lambda=0.987, F(2,57)=0.373, p=0.691, and no difference after being qualified by groups; Wilk’s Lambda=0.913, F(2,57)=2.712, p=0.075.

Neutrophils count tests showed significant variation over the study period; Wilk’s Lambda=0.83, F(2,59)=6.031, p=0.004, but there was no difference after being qualified by groups; Wilk’s Lambda=0.998, F(2,59)=0.047, p=0.954.

Lymph tests showed significant change over time; Wilk’s Lambda=0.83, F(2,54)=5.547, p=0.0.006, but showed no difference after being qualified by groups; Wilk’s Lambda=0.983, F(2,54)=0.456, p=0.636.

Urea tests showed no change over time; Wilk’s Lambda=0.968, F(2,44)=0.725, p=0.49; and no change after being qualified by groups; Wilk’s Lambda=0.968, F(2,44)=0.727, p=0.489.

In general, there was no significant difference between both treatment groups regardin Total Leukocyte Count, Neutrophils count, Lymph, Alanine transaminase, and Urea (Table 2).

### Secondary outcomes

In the S.L. group, there are 51 (41%) patients were achieved virological clearance within 15±5 days with median time 15 days, while in the OCH group, 54 (43%) patients were achieved virological clearance within 15±5 days with median time 15 days (Fig 5). There was no statistically significant difference between S.L. group and OCH group (CI: 0.257 – 0.489, p=0.76).

**Figure 5:** Kaplan-Meier plot for treatment groups considering the time to undetectable SARS-COV-2 RNA on two consecutive nasopharyngeal swabs.

Kaplan-Meier plot showed that the S.L. group and OCH group were no significantly regarding the virological clearance over time (p=0.76) (Fig 5).

## Discussion

COVID-19 is a global pandemic that has affected people all over the world. While most infected cases tend to be mild, some people develop respiratory problems that can lead to severe lung injury (15). To fight the current SARSCoV2 pandemic, which has resulted in COVID19, effective, powerful therapeutic strategies with minimal side effects are urgently needed (16). when there are no successful consolidated therapies available during epidemics, there is a tendency to use treatments based on preclinical study findings or observational trials with significant limitations (17). There are currently no known therapies for COVID-19, but many options are being considered, including experimental antivirals (15). Direct antiviral combination therapy such as Ledipasvir/Sofosbuvir demonstrated adequate Efficacy in treating HCV with a good safety profile that included minimal side effects and was well tolerated during treatment (18,19). Antiviral drugs that target particular viral targets are also the most successful way to stop the virus from spreading (20). This single-blinded randomized control study looked at antiviral drugs (Ledipasvir/Sofosbuvir) compared to standard treatment COVID-19. We summarized antiviral mechanism data and findings to subsidize decisions related to COVID-19 pharmacological therapy by providing clinically accessible evidence-based information in a clear interpretation. Also, Chen et al, (2020), recommended the drugs, Epclusa (sofosbuvir / velpatasvir) and Harvoni (sofosbuvir/ledipasvir) for managing COVID-19 infected patients owing to their dual inhibitory actions on two viral enzymes (21).

The previous studies (22–25) showed that Sofosbuvir / Daclatasvir regain had a faster time to recover from COVID-19 than Lopinavir/ritonavir, leading to the suggestion of using the Ledipasvir/Sofosbuvir combination as a better treatment choice than other direct antiviral agents in the management of COVID-19 (15,18,26). Wu et al. looked at several antiviral drugs, including favipiravir, oseltamivir, lopinavir, chloroquine, and hydroxychloroquine (12). Still, the efficacy of sofosbuvir alone should be tested as well. <u>Nourian</u> et al. mentioned that when Ledipasvir/Sofosbuvir was added to the standard of treatment, the clinical response time was shortened. However, there were no differences in clinical response rates, hospital and ICU stay lengths or 14-day mortality. There were no significant adverse events discovered. There are little data on the efficacy of antivirals against SARS-CoV2. If antiviral drugs are considered, it seems that they should be started as soon as possible during the early stages of the infection, when lung tissue damage has not progressed. Antiviral drugs cannot benefit once the inflammatory phase has begun and a cytokine storm has occurred. Sofosbuvir is a medication available in many countries and can treat mild to moderate COVID-19. However, larger sample sizes are required in clinical trials to confirm sofosbuvir’s efficacy in the treatment of COVID-19 (21). COVID-19 is currently being treated with symptomatic intensive intervention and supportive therapy (20). Although COVID-19 is most often associated with cough and fever (28), dyspnea, cough, and influenza-like illness are widespread side effects of sofosbuvir therapy (29,30).

In this single-blinded randomized control study, we found that the S.L. group was significantly higher than the OCH group to minimize the time to recover COVID-19 patients. There were no deaths in the S.L. group, but six deaths were in the OCH group. Seven patients in the S.L. group and six patients in the OCH group were admitted to ICU. Sofosbuvir shortened length of hospital stay was substantially compared to standard care time. Even though there were no deaths in the S.L group, Larger-scale experiments seem appropriate.

In general, there was no significant difference between both treatment groups regarding Total Leukocyte Count, Neutrophils count, Lymph, Alanine transaminase, and Urea.

Proportion of patients with undetectable SARS-COV-2 RNA on two consecutive nasopharyngeal swabs didn’t reach a statistical significance as detected by Kaplan–Meier curve through the treatment period, however, this doesn’t compromise the significant clinical outcomes for Sofosbuvir plus Ledipasvir treatment as Numerous studies have shown that identification of SARS-COV 2 RNA lasts longer than the resolution of COVID 19 symptoms and can continue for several weeks or months. (31) As regard the pneumonia recovery based on CT changes, this study revealed a non-significant increase of the CT stationary and progressive changes among SL patients at day 5, However, the increase in regressive changes among SL patients was significant at day 10. It is assumed that Sofosbuvir-Ledipasvir combination, with their potent antiviral effects, decreased the viral load, hence minimizing the pathologic impact of the virus on the lungs more than HCQ. This data is promising for further economic analysis longer follow up periods to evaluate long-term or permanent lung damage including fibrosis. (32)

## Conclusion

This single-blinded randomized controlled study looked at antiviral drugs (Ledipasvir/Sofosbuvir) compared to standard treatment for patients with moderate COVID-19 infection. We summarized antiviral mechanism data and findings to subsidize decisions related to COVID-19 pharmacological therapy by providing clinically accessible evidence-based information in a clear interpretation. We found that the S.L. group was significantly higher than the OCH group to minimize the time to recover COVID-19 patients. There were no deaths in the S.L. group, but six deaths were in the OCH group. Seven patients in the S.L. group and six patients in the OCH group were admitted to ICU. Sofosbuvir shortened length of hospital stay was substantially compared to standard care time. Even though there were no deaths in the S.L group, Larger-scale experiments seem appropriate.

## Supporting information

D:\papers\SL

D:\papers\SL

## Data Availability

Data Availability from the Corresponding Author

