## Supplementary material for "Efficacy of Sofosbuvir plus Ledipasvir in Egyptian patients with COVID-19 compared to standard treatment: Randomized controlled trial": D:\papers\SL

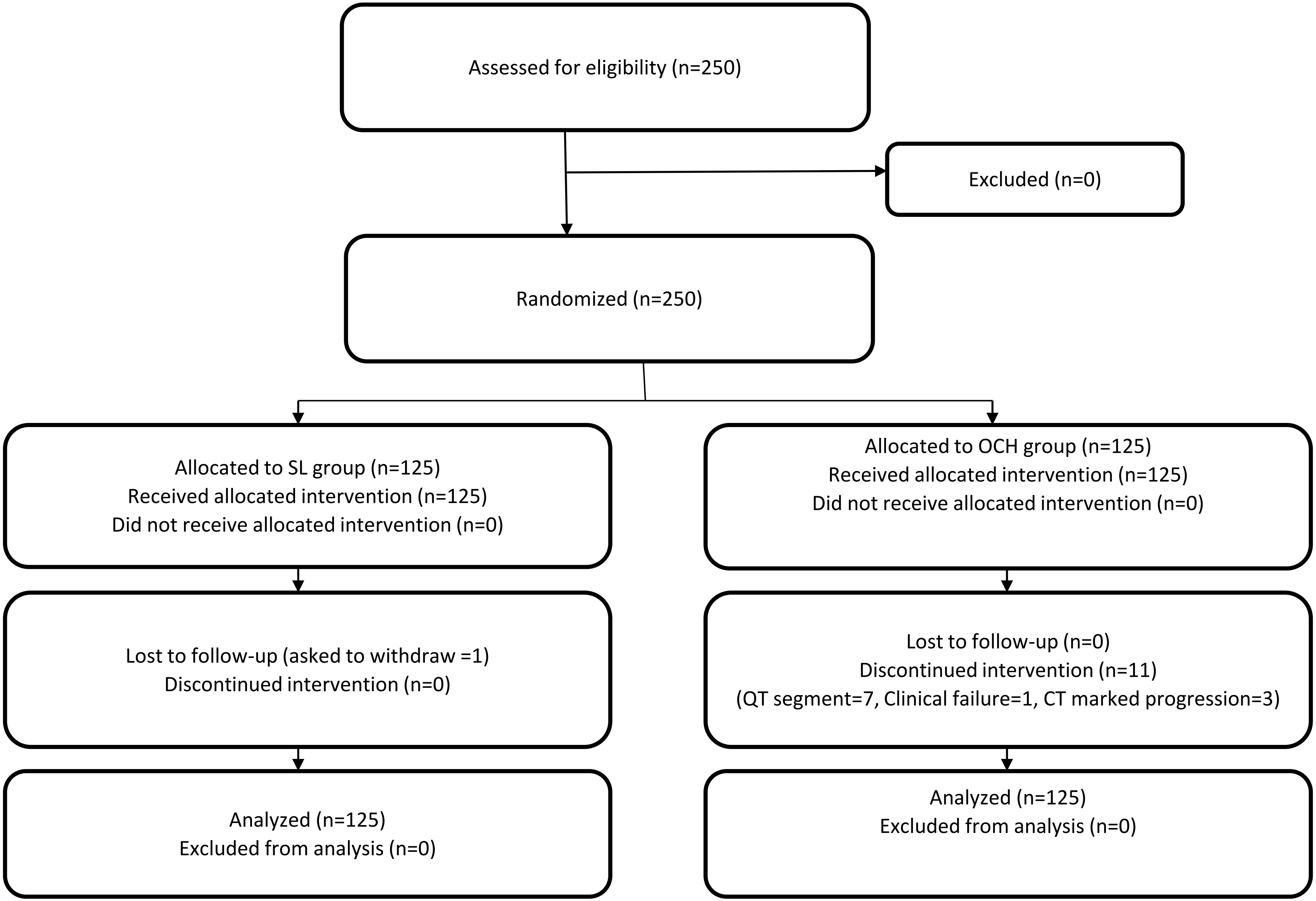


**Figure 1:** Study flow diagram

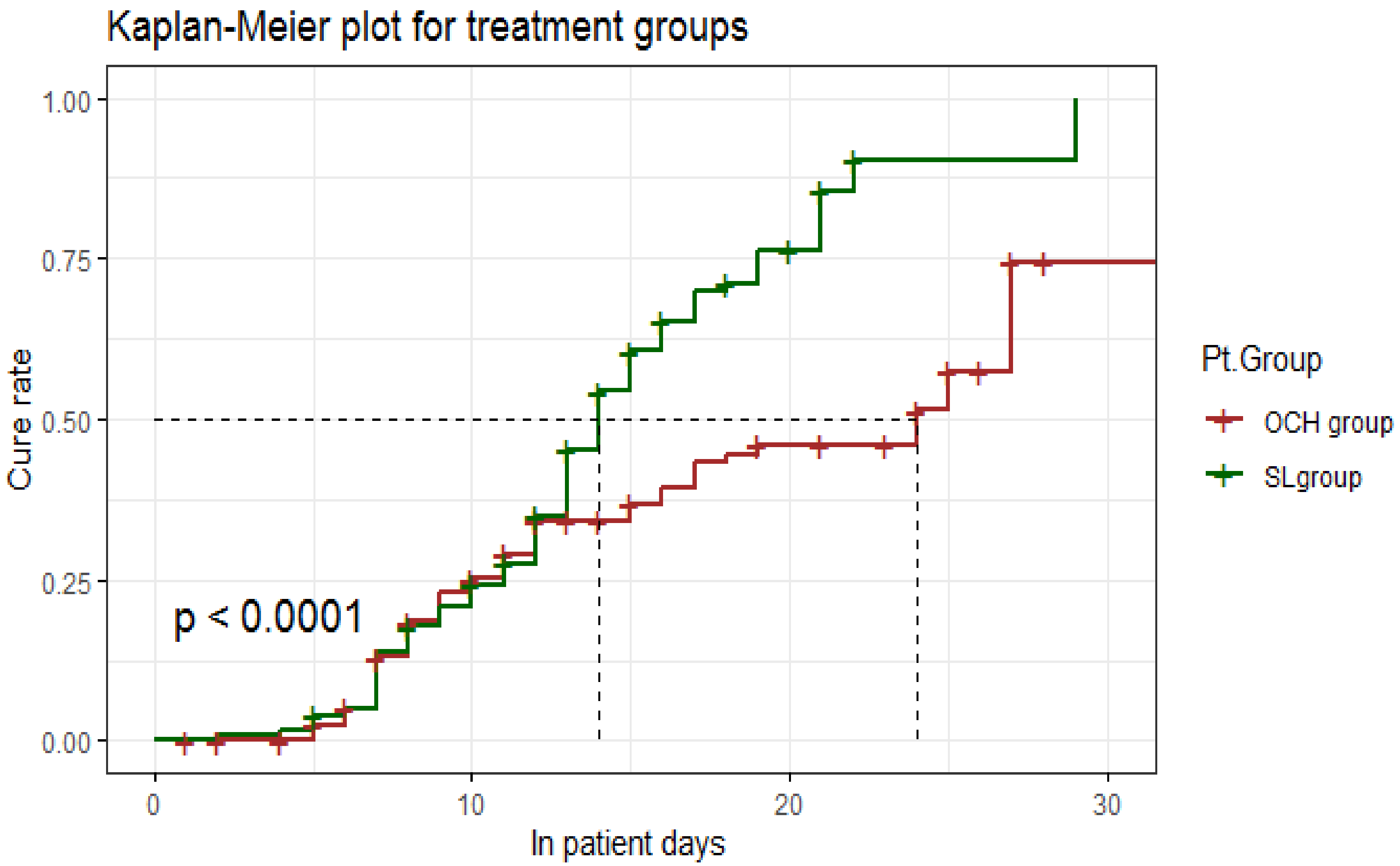


**Figure 2:** Kaplan-Meier plot for treatment groups time to clinical cure (log-rank=16.98).

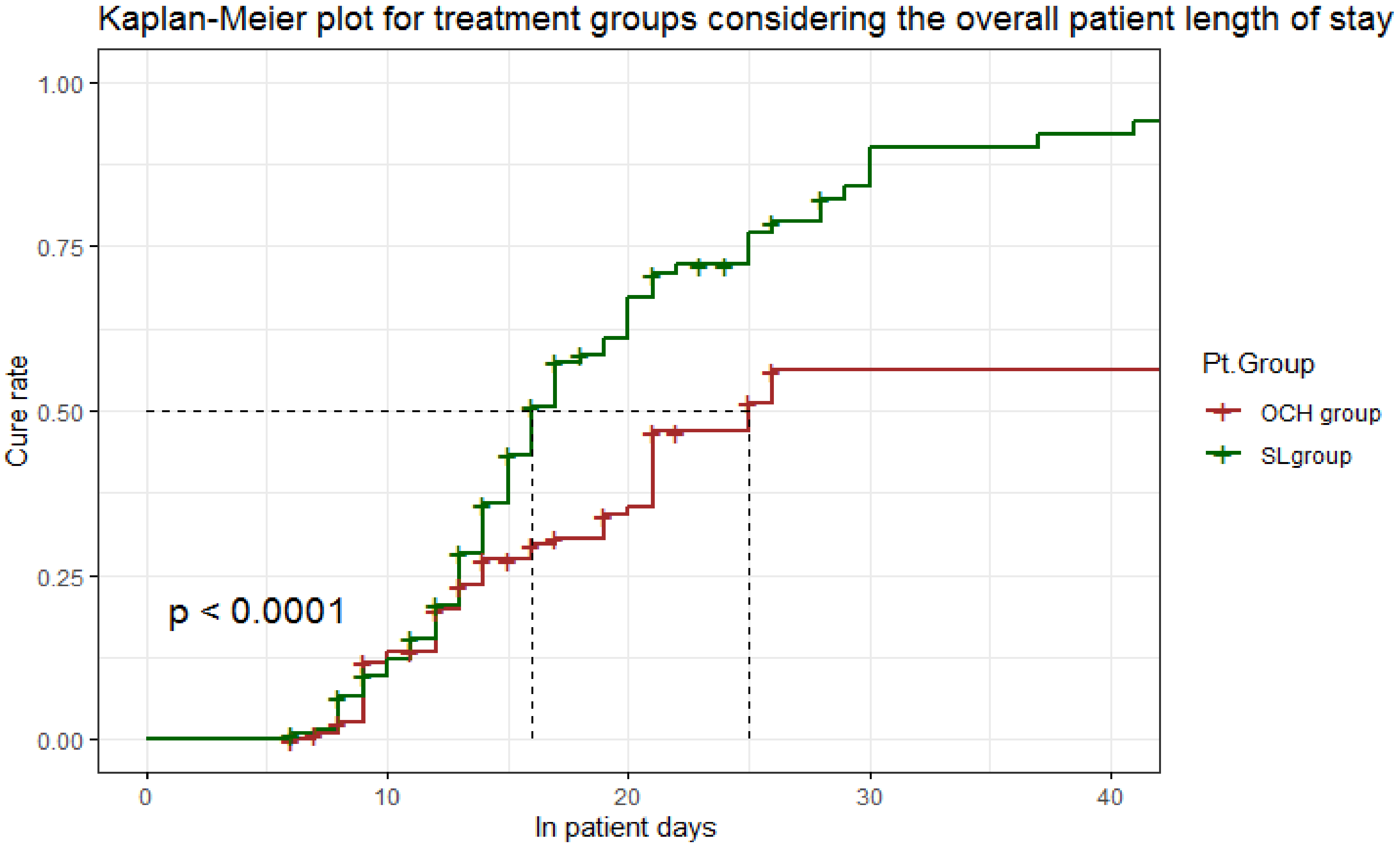


**Figure 3:** Kaplan-Meier plot for treatment groups considering the overall patient length of stay.

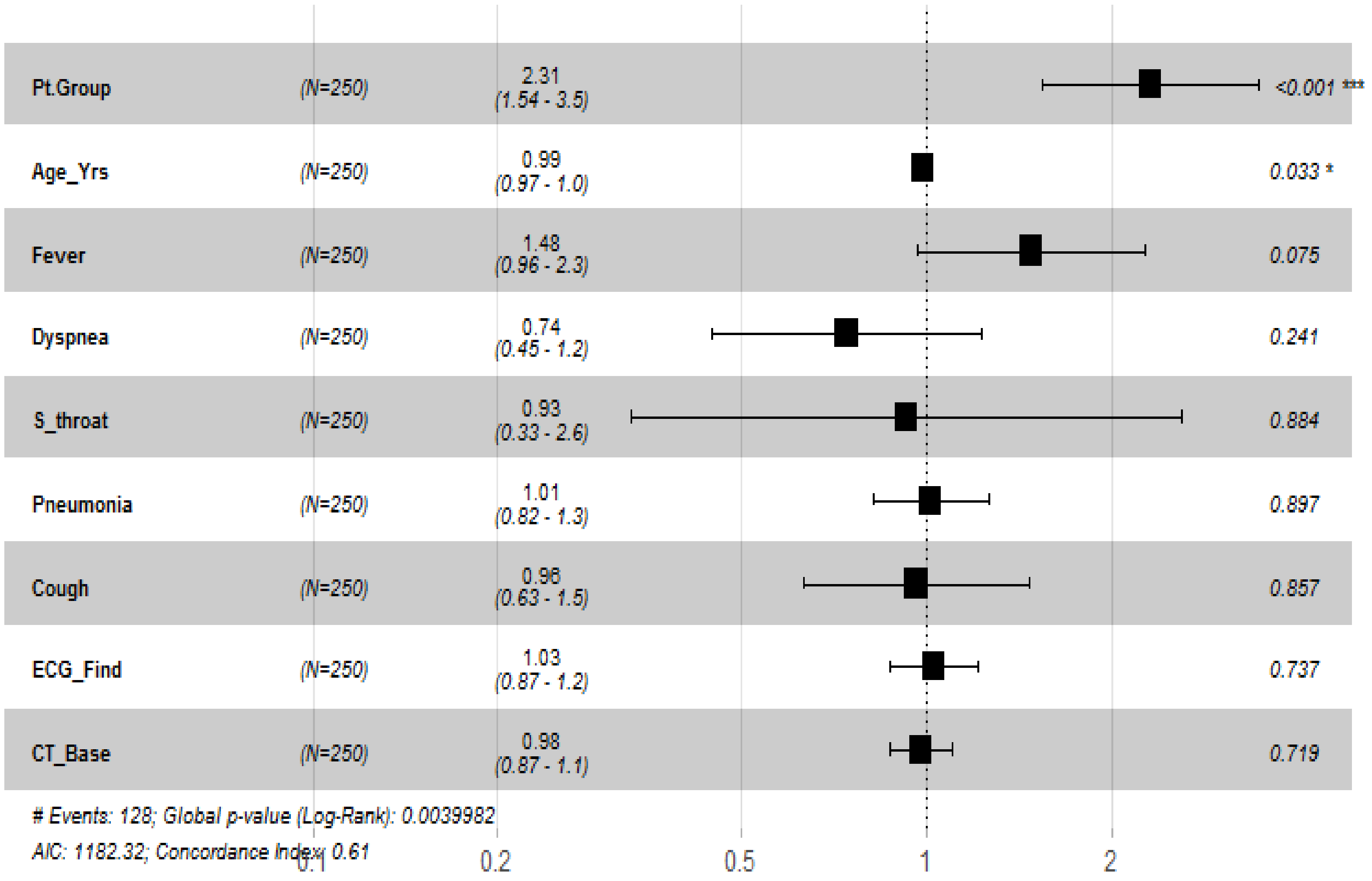


**Figure 4:** Cox regression between the treatments groups regarding suspected covariates at baseline.

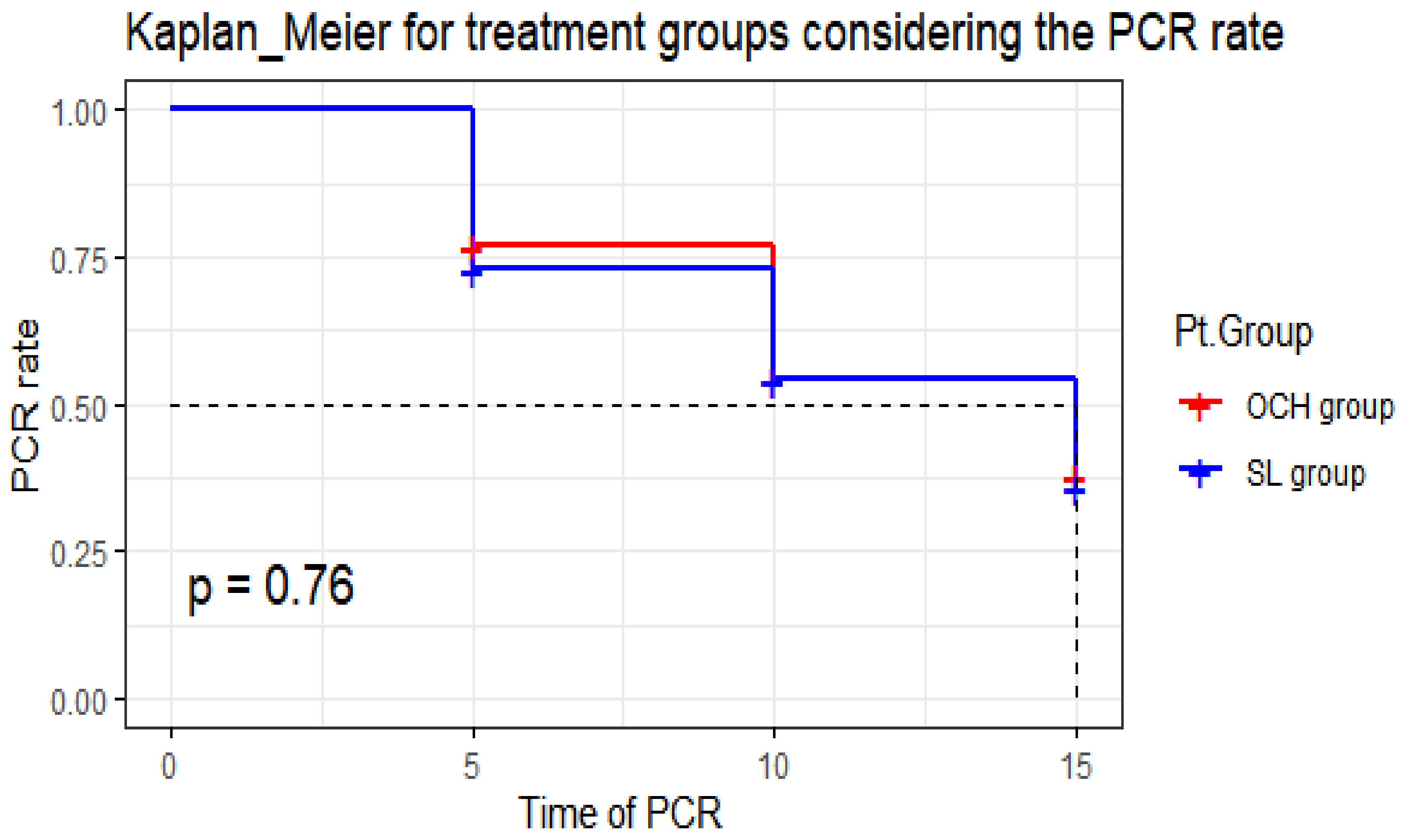


**Figure 5:** Kaplan-Meier plot for treatment groups considering the time to undetectable SARS-COV-2 RNA on two consecutive nasopharyngeal swabs.
