## Supplementary material for "Efficacy of Sofosbuvir plus Ledipasvir in Egyptian patients with COVID-19 compared to standard treatment: Randomized controlled trial": D:\papers\SL

**Table 1**: Baseline characteristics of the participants compared by independent t test. SL group: Sofosbuvir plus Ledipasvir, OCH group: Oseltamivir plus Hydroxychloroquine in combination with Azithromycin. Data were represented as mean ± SD or frequency (percentage). NI: no information

| Parameter | | S.L. group (n=125) | OCH group (n=125) | p-value |
|---|---|---|---|---|
| Age | | 46.83 ± 15.24 | 40.24 ± 14.7 | 0.001 |
| Gender | Male | 1 (0.8%) | 0 (0%) | 0.113 |
| | Female | 89 (71.2%) | 102 (81.6%) | |
| **Clinical and radiological findings** | | | | |
| Fever | | 71 (56.8%) | 75 (60.0%) | 1 |
| Sore Throat | | 3 (2.4%) | 5 (4.0%) | 1 |
| Dyspnea | | 21 (16.8%) | 21 (16.8%) | 1 |
| Cough | | 69 (55.2%) | 59 (47.2%) | 0.071 |
| Pneumonia | | 90 (72.0%) | 58 (46.4%) | 0.003 |
| ECG abnormal | | 29 (23.2%) | 35 (28.0%) | 0.591 |
| ECG Findings | Normal | 90 (72.0%) | 49 (39.2%) | <0.001 |
| | T Wave | 7 (5.6%) | 3 (2.4%) | |
| | QT Segment | 6 (4.8%) | 13 (10.4%) | |
| | Other | 16 (12.8%) | 19 (15.2%) | |
| CT Chest | Scattered Opacities | 60 (48.0%) | 48 (38.4%) | <0.001 |
| | Consolidated Patches | 5 (4.0%) | 6 (4.8%) | |
| | GG appearance | 37 (29.6%) | 65 (52.0%) | |
| | Unremarkable | 0 (0.0%) | 0 (0.0%) | |
| | Lower Lobe Pneumonia Patch | 10 (8.0%) | 6 (4.8%) | |
| | Scattered Pneumonia | 13 (10.4%) | 0 (0.0%) | |
| **Laboratory findings** | | | | |
| Total Leukocyte Count | | 5.92 ± 5.68 | 5.71 ± 2.37 | 0.720 |
| Neutrophils count | | 60.28 ± 13.00 | 58.69 ± 14.94 | 0.384 |
| Lymph | | 33.87 ± 10.93 | 35.80 ± 14.15 | 0.239 |
| Neutrophil lymphocyte ratio | | NI | 204.61 ± 73.68 | - |
| Platelet count | | 209.29 ± 85.68 | NI | - |
| Alanine transaminase | | 35.88 ± 29.19 | 29.62 ± 13.28 | 0.041 |
| Urea | | 33.27 ± 14.88 | 1.13 ± 0.31 | <0.001 |
| Creatinine | | 1.15 ± 0.43 | 30.38 ± 19.82 | <0.001 |
| C-reactive protein | | NI | 318.18 ± 331.22 | - |
| D-dimer | | 357.75 ± 443.92 | 136.33 ± 140.27 | 0.019 |
| Serum ferritin | | 386.37 ± 510.42 | 195.80 ± 323.49 | 0.110 |
| Thyroglobulin | | 186.13 ± 102.17 | 219.39 ± 92.63 | 0.177 |
| Lactate dehydrogenase | | 255.76 ± 90.90 | 70.80 ± 132.85 | <0.001 |
| Fibrinogen | | 343.14 ± 84.16 | 261.60 ± 123.65 | 0.152 |

**Table 2:** Repeated measures ANOVA showing the change over time of the laboratory findings.

| Variable | Within groups | | | | | | Between groups |
|---|---|---|---|---|---|---|---|
| | S.L. Group | | | OCH Group | | | |
| | pre | post | p-value | pre | Post | p-value | P-value |
| Total Leukocyte Count | 6.63 ± 10.14 | 6.85 ± 3.44 | 0.319 | 5.92 ± 2.21 | 7.19 ± 3.7 | 0.016 | 0.075 |
| Neutrophils count | 61.54 ± 14.92 | 57.59 ± 10.93 | 0.037 | 61.05 ± 14.03 | 58.58 ± 12.9 | 0.079 | 0.954 |
| Lymph | 33.33 ± 11.48 | 35.75 ± 10.53 | 0.166 | 35.91 ± 13.87 | 38.62 ± 13.88 | 0.043 | 0.636 |
| Urea | 37.78 ± 19.18 | 35.81 ± 13.66 | 0.229 | 0.98 ± 0.31 | 1.01 ± 0.29 | 0.859 | 0.489 |

Table (3): Comparison between the radiological changes based on CT findings at the three study points

| | | OCH | SL | p-value |
|---|---|---|---|---|
| CT day5 | Progressive | 19 (15.2%) | 24 (19.4%) | 0.244 |
| | Regressive | 42 (33.6%) | 42 (33.9%) | |
| | Stationary | 40 (32.0%) | 46 (37.1%) | |
| CT day10 | Progressive | 5 (4.0%) | 6 (4.8%) | 0.033 |
| | Regressive | 24 (19.2%) | 44 (35.5%) | |
| | Stationary | 27 (21.6%) | 26 (21.0%) | |
